# Probing the Depths for Diagnostic Performance of Biparametric and Multiparametric MRI for Prostate Cancer Detection- A Meta-Analysis

**DOI:** 10.1101/2024.02.12.24302703

**Authors:** Dev Desai, Vismit Gami, Abhijay B. Shah, Dwija Raval, Parth Gupta, Hetvi Shah

## Abstract

**Background:** Prostate cancer is a malignancy that originates in the prostate gland,and can vary in aggressiveness, often requiring a combination of diagnostic methods and imaging for accurate detection and management. In an attempt to ensure timely diagnosis and prevent complications, the choice of the right diagnostic modality becomes crucial. MRI has gained prominence in the diagnosis of prostate cancer. MRI aids prostate cancer diagnosis by pinpointing suspicious areas for in-depth investigation and guiding precise biopsies, enhancing accuracy. It also informs treatment plans by visualizing tumour extent and assists in monitoring disease progression during active surveillance. The purpose of this meta-analysis is to ascertain the accuracy of MRI in diagnosing prostate cancer.

**Methodology:** Medical literature was comprehensively searched and reviewed without restrictions to particular study designs, or publication dates using PubMed, Cochrane Library, and Google Scholar databases for all relevant literature. The extraction of necessary data proceeded after specific inclusion and exclusion criteria were applied. In this Meta-Analysis, A total of 47 RCTs with 13,211 subjects were selected for comparing Multiparametric MRI vs. Gold Standard and A total of 23 RCTs with 3440 subjects were selected for comparing Biparametric MRI vs Gold Standard.

Two writers independently assessed the calibre of each study as well as the use of the Cochrane tool for bias risk apprehension. The statistical software packages RevMan (Review Manager, version 5.3), SPSS (Statistical Package for the Social Sciences, version 20), and Excel in Stata 14 were used to perform the statistical analyses.

**Results:** We calculated the sensitivity and specificity of multiparametric as well as biparametric MRI. The Multiparametric MRI demonstrates a sensitivity of 0.84 (95% confidence interval: 0.83 - 0.85) and specificity of 0.69 (95% confidence interval: 0.68 - 0.70). Meanwhile, Biparametric MRI shows a sensitivity of 0.85 (95% confidence interval: 0.84 - 0.86) and specificity of 0.71 (95% confidence interval: 0.69 - 0.73).

**Conclusion:** In conclusion, both Multiparametric MRI and Biparametric MRI exhibit high sensitivity values of 0.84 and 0.85, respectively, indicating their ability to accurately detect prostate cancer. MRI is a robust diagnostic tool for prostate cancer due to its high-resolution imaging and multiparametric approach. It enables targeted biopsies, informs treatment plans, and aids in active surveillance.

## Introduction

Prostate cancer is the alternate leading cause of cancer death in American men, behind only lung cancer. Roughly 1 out of every 8 men will receive a prostate cancer diagnosis at some point in their lives, highlighting the need for effective approaches to screening, diagnosing, and treating this condition.

Prostate cancer is more likely to develop in aged men and in non-Hispanic Black men. About 6 cases in 10 are diagnosed in men who are 65 years or above, and relatively rare in men under the age of 40. The prostate cancer death rate declined by about half from 1993 to 2013, probably due to earlier discoveries and advances in treatment[1].

Prostate-specific antigen (PSA) testing and digital rectal examination (DRE), are frequently used in clinical practice and are endorsed as early detection methods for the disease. Although neither of the two tests are suitable for use in isolation for prostate cancer screening.

Diagnosis in men at risk, or those with elevated PSA levels or abnormal digital rectal examinations, is done via TRUS-guided biopsy. Eight to twelve biopsy cores are used to sample various prostatic regions[2]. Being an invasive procedure, it brings significant discomfort to patients.

The Gleason grading system describes the unusual features of prostate cancer cells obtained after biopsy and their likelihood of spread. A lower Gleason score indicates a less severe and less prolific malignancy whereas, higher values represent a more aggressive tumour. Grade 3 represents the minimal evaluation for a tumour, with grades below 3 indicating normal or nearly normal cells. Most cancers are assigned a Gleason score of 6 (consisting of 3+3) or 7 (comprising 3+4 or 4+3), which results from adding the two most common grades together [3]. MRI-guided biopsies, often known as targeted biopsies, have been a fast-expanding area of clinical research in the discipline of urologic oncology. Compared with traditional template biopsies, MRI-targeted biopsies were found to be equivalent for the diagnosis of prostate cancer and have a lower prevalence of clinically inconsequential cancer detection.[4]

Multiparametric prostate MRI (MP-MRI) has evolved into a precise structural and clinical imaging method for detecting, locating, and staging prostate cancer [7–9]. As per Prostate Imaging Reporting and Data System version 2 (PI-RADS V2) guidelines, MP-MRI comprises T2-weighted (T2W), diffusion-weighted (DWI), dynamic contrast-enhanced (DCE), and occasionally MR spectroscopy[5]. Conversely, biparametric MRI (bpMRI) employs T2-weighted images and DWI, excluding DCE and/or MR spectroscopy from the imaging protocol.

## Prisma flowchart

**Figure 1 :**
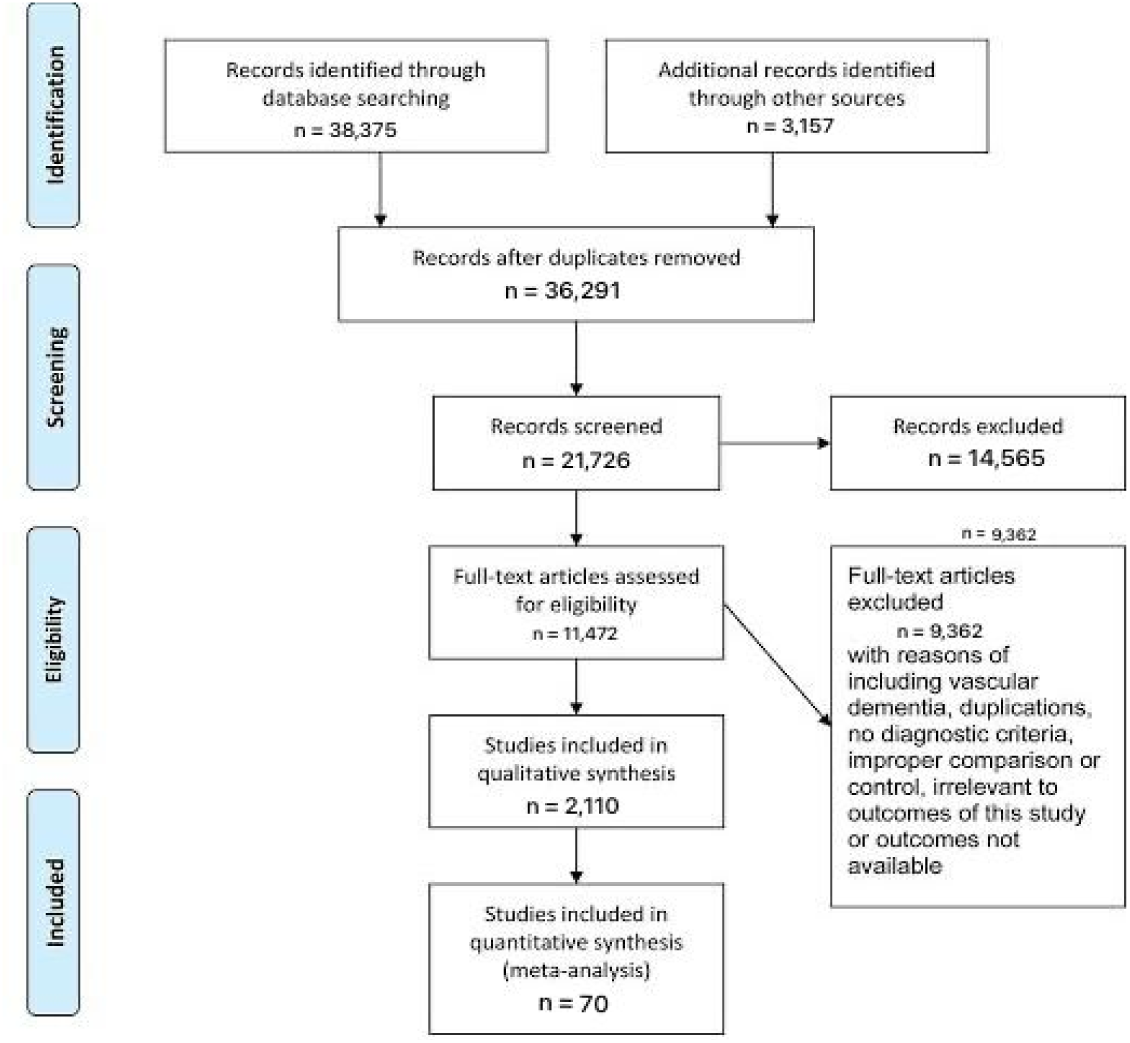
PRISMA FLOWCHART.

## Methodology

### Data Collection

For the collection of the data, a search was done by two individuals using PubMed, Google Scholar, and Cochrane Library databases for all relevant literature. Full - Text Articles written only in English were considered.

The medical subject headings (MeSH) and keywords ‘Biparametric MRI’, ‘Multiparametric MRI’, and ‘Prostate cancer’ were used. References, reviews, and meta-analyses were scanned for additional articles[6].

### Inclusion And Exclusion Criteria

Titles and abstracts were screened, and Duplicates and citations were removed. References of relevant papers were reviewed for possible additional articles. Papers with detailed patient information and statically supported results were selected.We searched for papers that show more accurate diagnoses, where modalities considered were Biparametric and Multiparametric MRI for detection of prostate cancer.

The inclusion criteria were as follows: (1) studies that provided information about the accurate diagnosis with Biparametric and Multiparametric MRI; (2) studies published in English; (3) Studies comparing Biparametric MRI with Multiparametric MRI as well as comparisons to the Gold Standard (biopsy) as a Diagnosis modality for cases of prostate carcinoma.

The exclusion criteria were: (1) articles that were not full text, (2) unpublished articles, and (3) articles in other languages.

### Data Extraction

Each qualifying paper was independently evaluated by two reviewers. Each article was analyzed for the number of patients, their age, procedure modality, and incidence of the pre decided complications. Further discussion or consultation with the author and a third party was used to resolve conflicts. The study’s quality was assessed using the modified Jadad score. In the end, According to PRISMA, a total of 47 RCTs pertaining to use of Multiparametric MRI with a total of 13,211 patients along with 23 RCTs pertaining to Biparametric MRI with a total of 3440 patients were selected for further analysis.

### Assessment Of Study Quality

Using the QualSyst tool, two writers independently assessed the caliber of each included study. This test consists of 10 questions, each with a score between 0 and 2, with 20 being the maximum possible overall score. Two authors rated each article independently based on the above criteria. The interobserver agreement for study selection was determined using the weighted Cohen’s kappa (K) coefficient. For deciding the bias risk for RCTs, we also employed the Cochrane tool. No assumptions were made about any missing or unclear information. there was no funding involved in collecting or reviewing data.

### Statistical Analysis

**t**he statistical software packages RevMan (Review Manager, version 5.3), SPSS (Statistical Package for the Social Sciences, version 20), Google Sheets, and Excel in Stata 14 were used to perform the statistical analyses. The data was obtained and entered into analytic software [21]. Fixed- or random-effects models were used to estimate Sensitivity, Specificity, positive predictive value (PPV), diagnostic odds ratios (DOR), and relative risk (RR) with 95 percent confidence intervals to examine critical clinical outcomes (CIs). Diagnosis accuracy and younden index were calculated for each result. Individual study sensitivity and specificity were plotted on Forest plots and in the receiver operating characteristic (ROC) curve. The forest plot and Fagan’s Nomogram were used to illustrate the sensitivity and specificity of different papers.

### Bias Study

The risk of bias was evaluated by using QUADAS-2 analysis. This tool includes 4 domains as Patient selection, Index test, Reference standard, Flow of the patients, and Timing of the Index tests

## Results

**Figure 2A :**
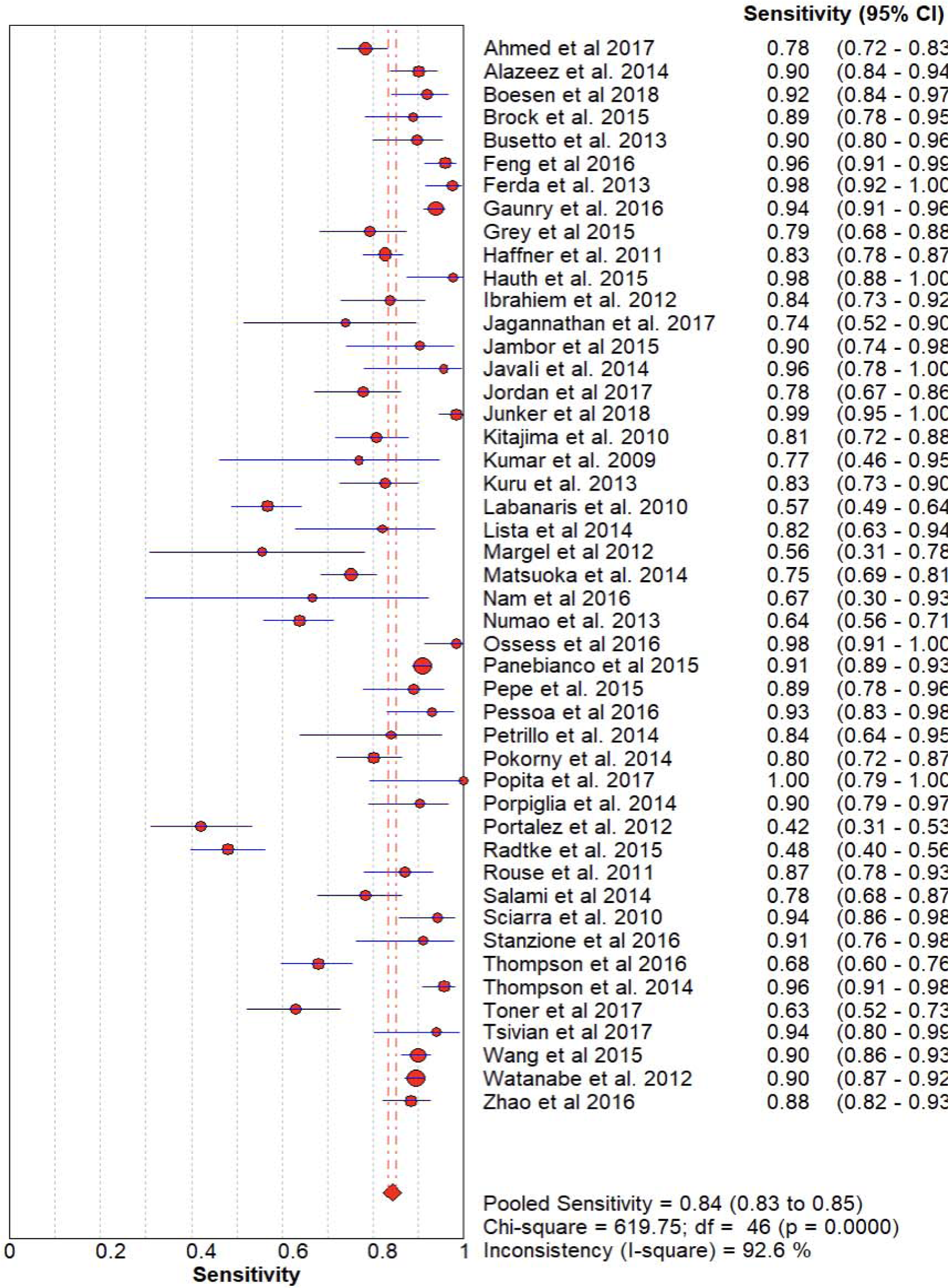
The forest chart summary for pooled sensitivity values of Multiparametric MRI.

**Figure 2B :**
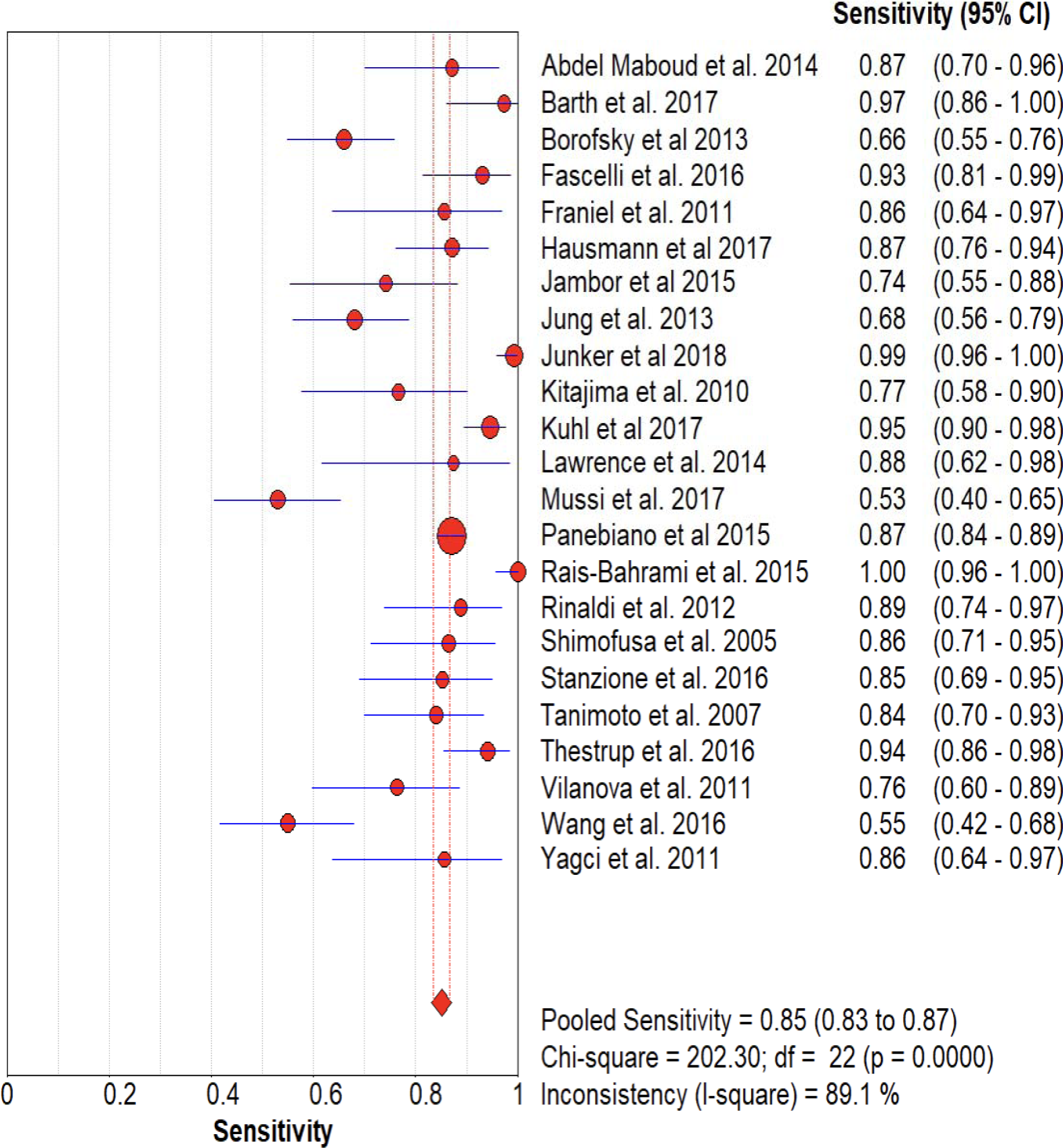
The forest chart summary for pooled sensitivity values for Biparametric MRI.

**Figure 3A :**
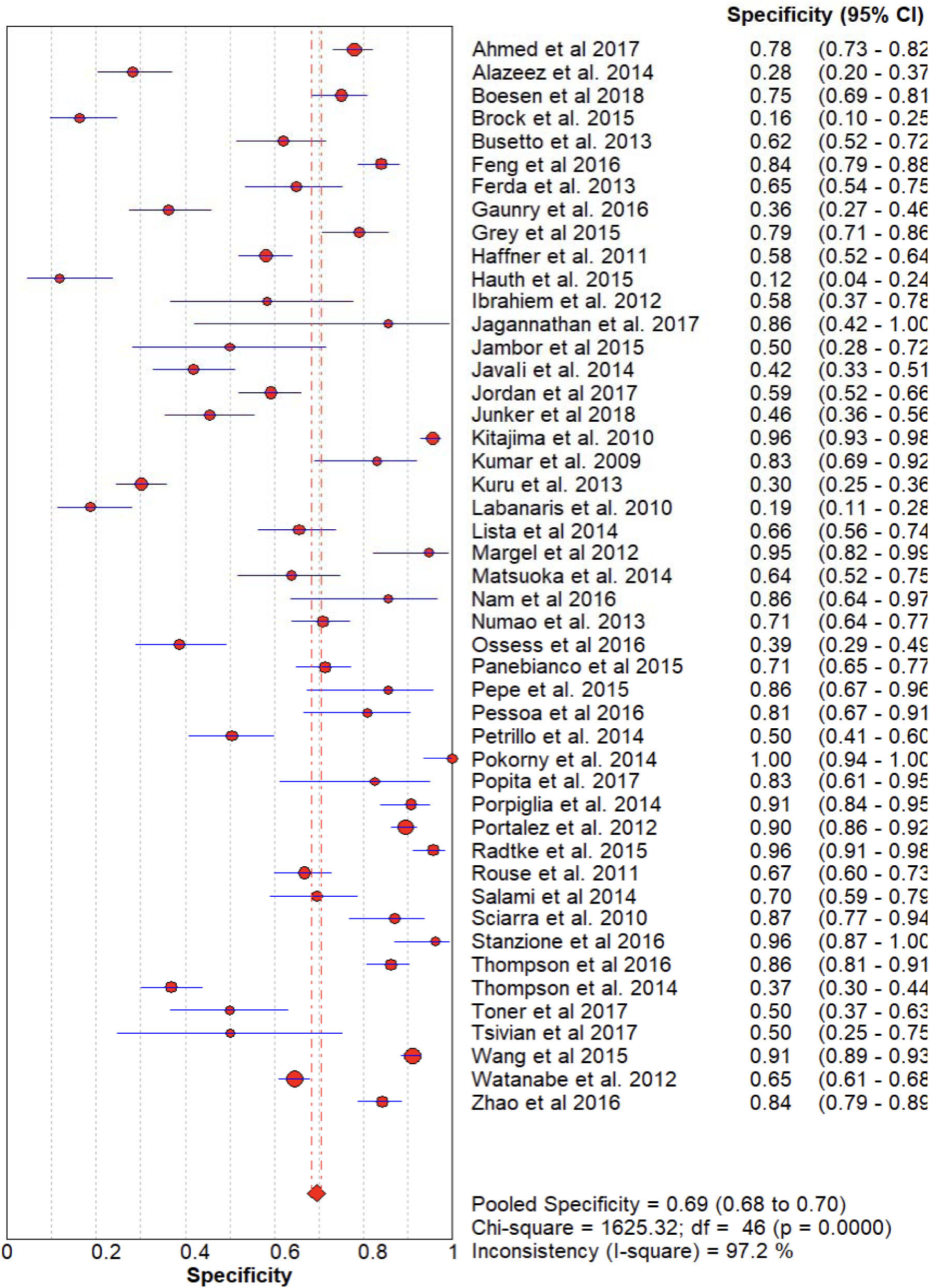
The forest chart summary for pooled specificity values for Multiparametric MRI.

**Figure 3B :**
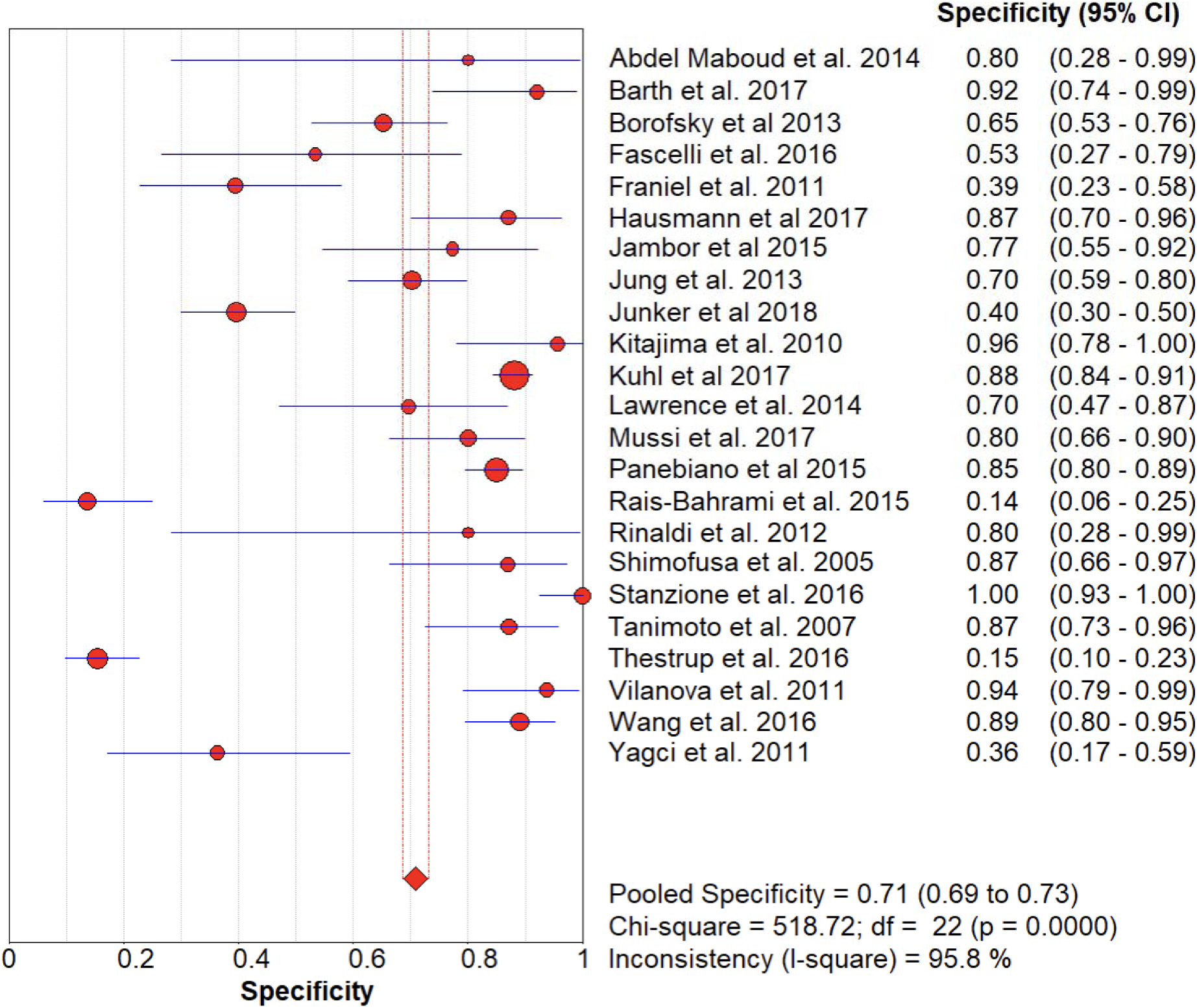
The forest chart summary for pooled specificity values for Biparametric MRI.

**Figure 4A :**
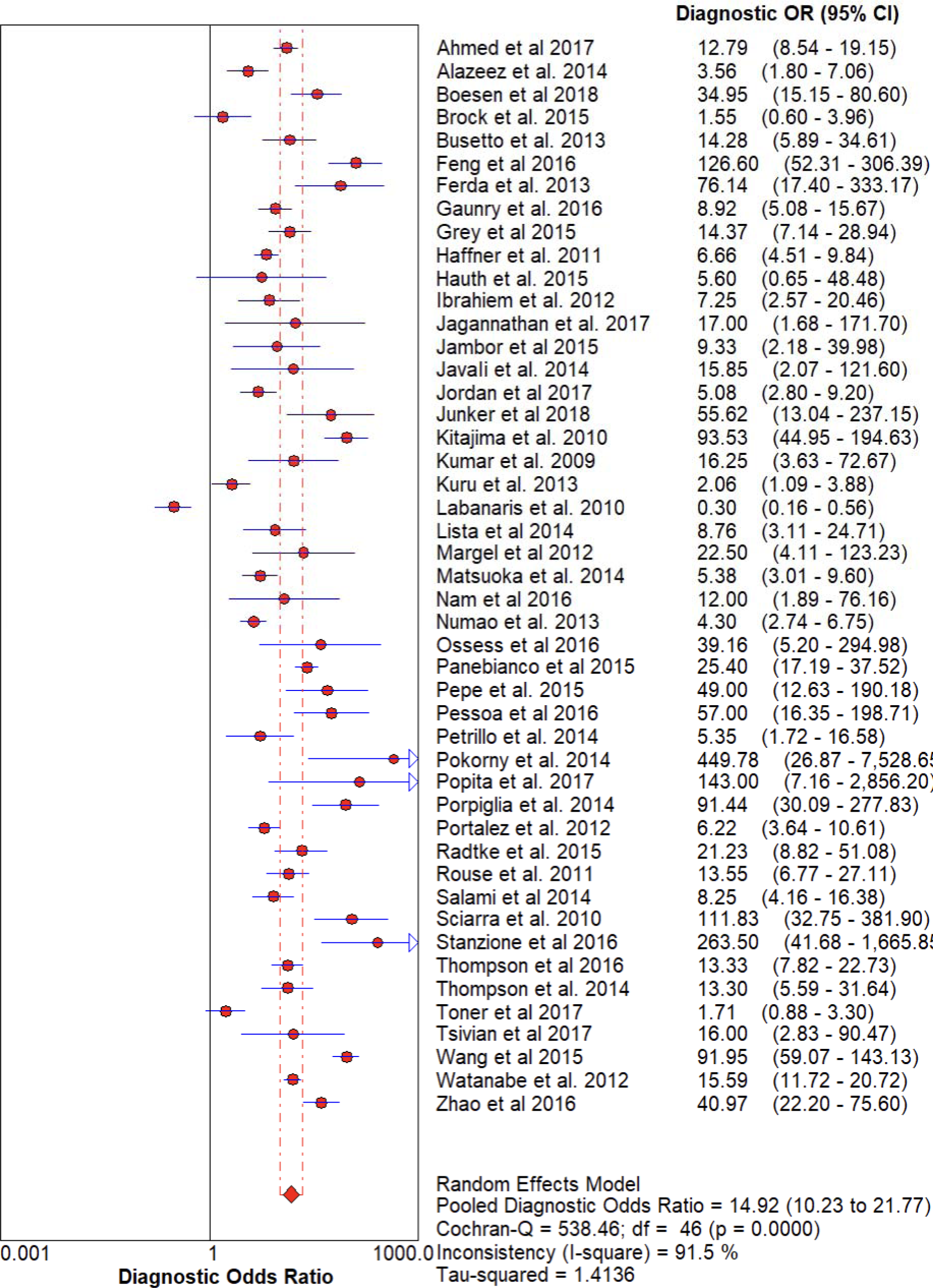
The forest chart summary for pooled Diagnostic Odds Ratio for Multiparametric MRI.

**Figure 4B :**
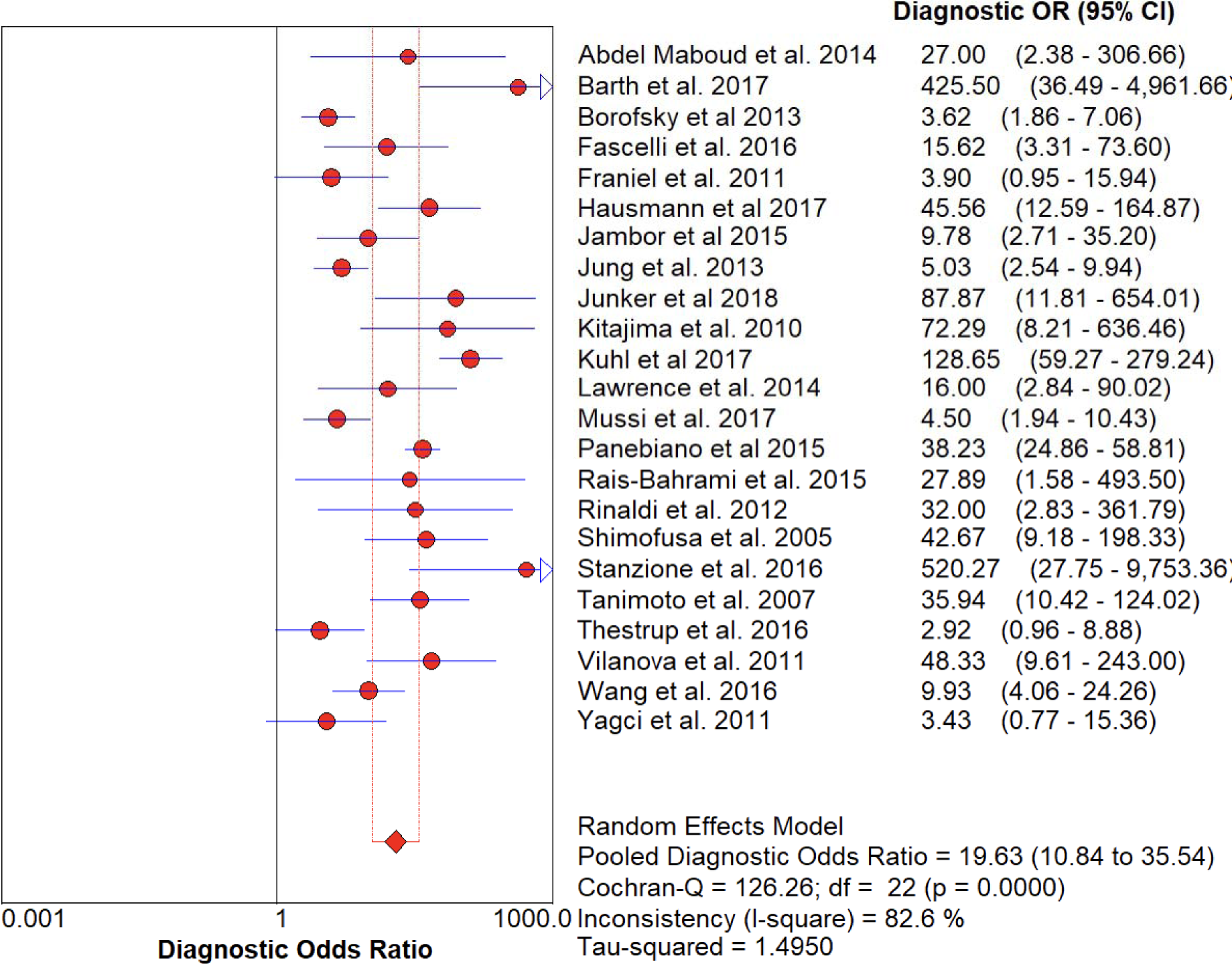
The forest chart summary for pooled Diagnostic Odds Ratio for Biparametric MRI.

**Figure 5A :**
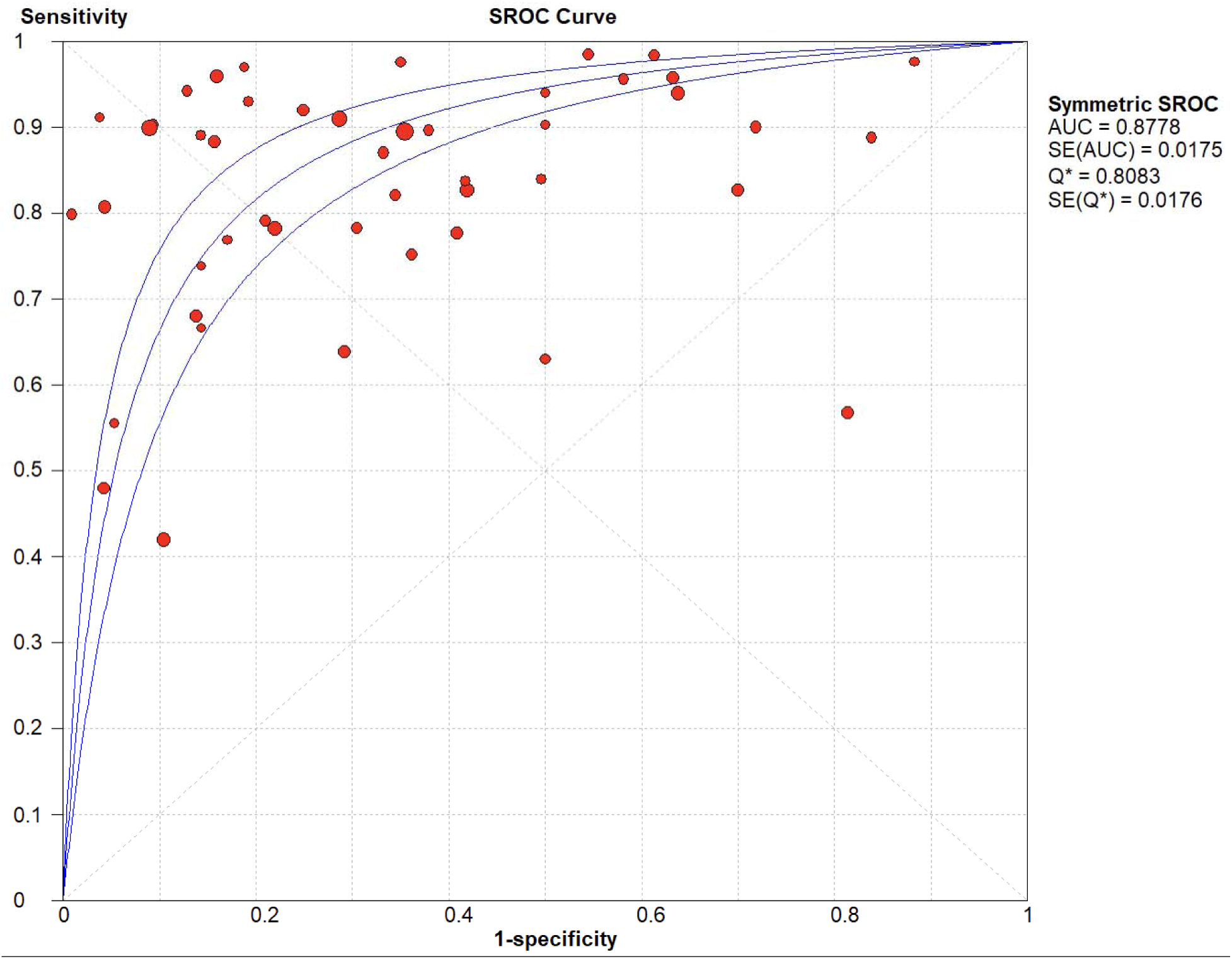
The SROC plot summary for Multiparametric MRI for Prostate Cancer.

**Figure 5B :**
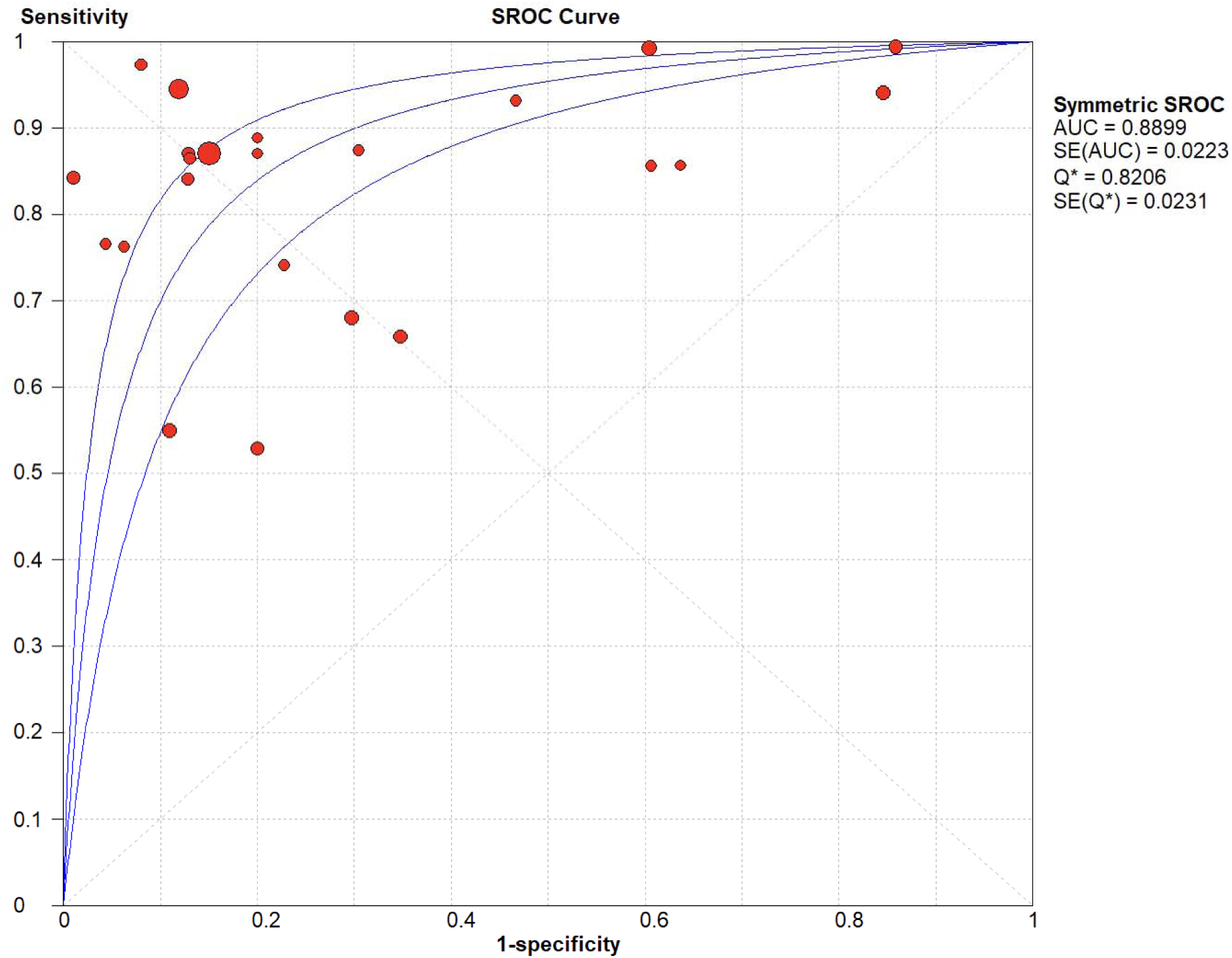
The SROC plot summary for Biparametric for Prostate Cancer.

**Figure 6A :**
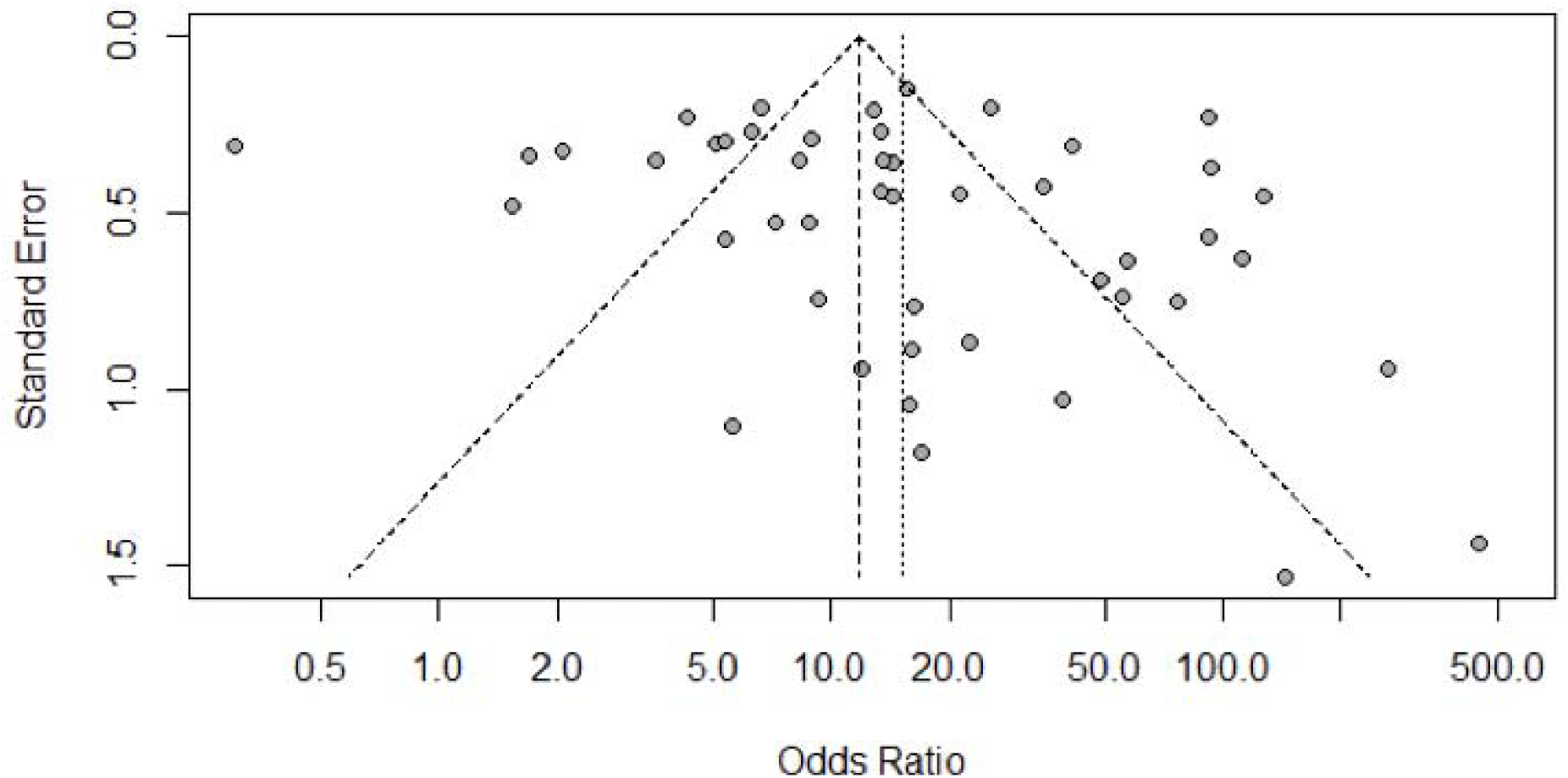
Funnel Plot for Multiparametric MRI for Prostate Cancer.

**Figure 6B :**
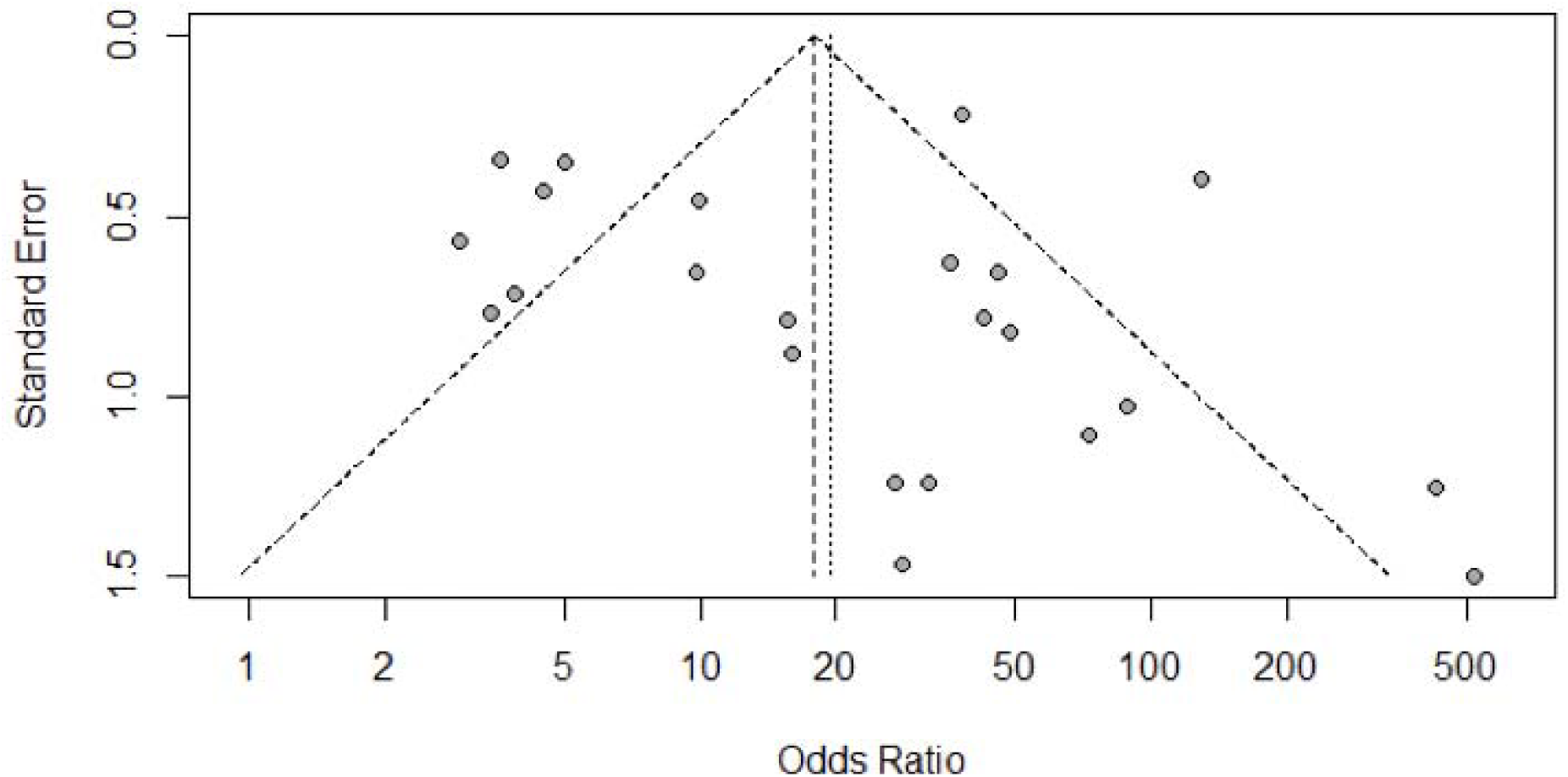
Funnel plot for Biparametric MRI for Prostate Cancer.

## Multiparametric MRI vs Gold Standard

Here, Table 1 describes all the descriptions of papers used for the study regarding the use of Multiparametric MRI in the diagnosis of Prostate Cancer. In Figure 2A, the Pooled Sensitivity Values for all papers being considered can be observed and compared amongst each other, while Figure 3A serves the same purpose in the context of Pooled Specificity Values. Figure 4A denotes the pooled Diagnostic Odds Ratio for the application of Multiparametric MRI. The same is illustrated in the SROC curve. (Figure 5A). Figure 6A represents Deek’s Funnel Plot. A total of 47 RCTs with 13,211 subjects were selected for the study, out of which 8 studies showed sensitivity above or equal to 95%, and 5 studies showed specificity above 95%. The value of True Positive (TP) was 5036, that of True Negative (TN) was 5024, that of False Positive (FP) was 2214, and that of False Negative (FN) was 937. With a confidence interval 95%, sensitivity, specificity and positive predictive values were calculated. The sensitivity of Multiparametric MRI is 0.84, with a CI of 95% in a range of 0.83 to 0.85. The specificity of Multiparametric MRI is 0.69, with a CI of 95% in a range of 0.68 to 0.70.

Figure 5A shows the summary of the ROC curve. It shows that the area under the curve for Multiparametric MRI was 0.8778 and the overall diagnostic odds ratio (DOR) was 14.92 with Younden Index being 0.55.

## Biparametric MRI vs Gold Standard

Here, Table 2 describes all the descriptions of papers used for the study regarding the use of Biparametric MRI in the diagnosis of Prostate Cancer. In Figure 2B, the Pooled Sensitivity Values for all papers being considered can be observed and compared amongst each other, while Figure 3B serves the same purpose in the context of Pooled Specificity Values. Figure 4B denotes the pooled Diagnostic Odds Ratio for the application of biparametric MRI. The same is illustrated in the SROC curve. (Figure 5B). Figure 6B represents Deek’s Funnel Plot. A total of 23 RCTs with 3440 subjects were selected for the study, out of which 4 studies showed sensitivity above or equal to 95%, and 2 studies showed specificity above 95%. The value of True Positive (TP) was 1623, that of True Negative (TN) was 1087, that of False Positive (FP) was 446, and that of False Negative (FN) was 284. With a confidence interval 95%, sensitivity, specificity and positive predictive values were calculated. The sensitivity of Multiparametric MRI is 0.85, with a CI of 95% in a range of 0.84 to 0.86. The specificity of Biparametric MRI is 0.71, with a CI of 95% in a range of 0.69 to 0.73.

<u>**Figure 5A shows the summary of the ROC curve. It shows that the area under the curve for Biparametric RI was 0.8899 and the overall diagnostic odds ratio (DOR) was 19.63 with Younden Index being 0.56.**</u>

**Figure 7A :**
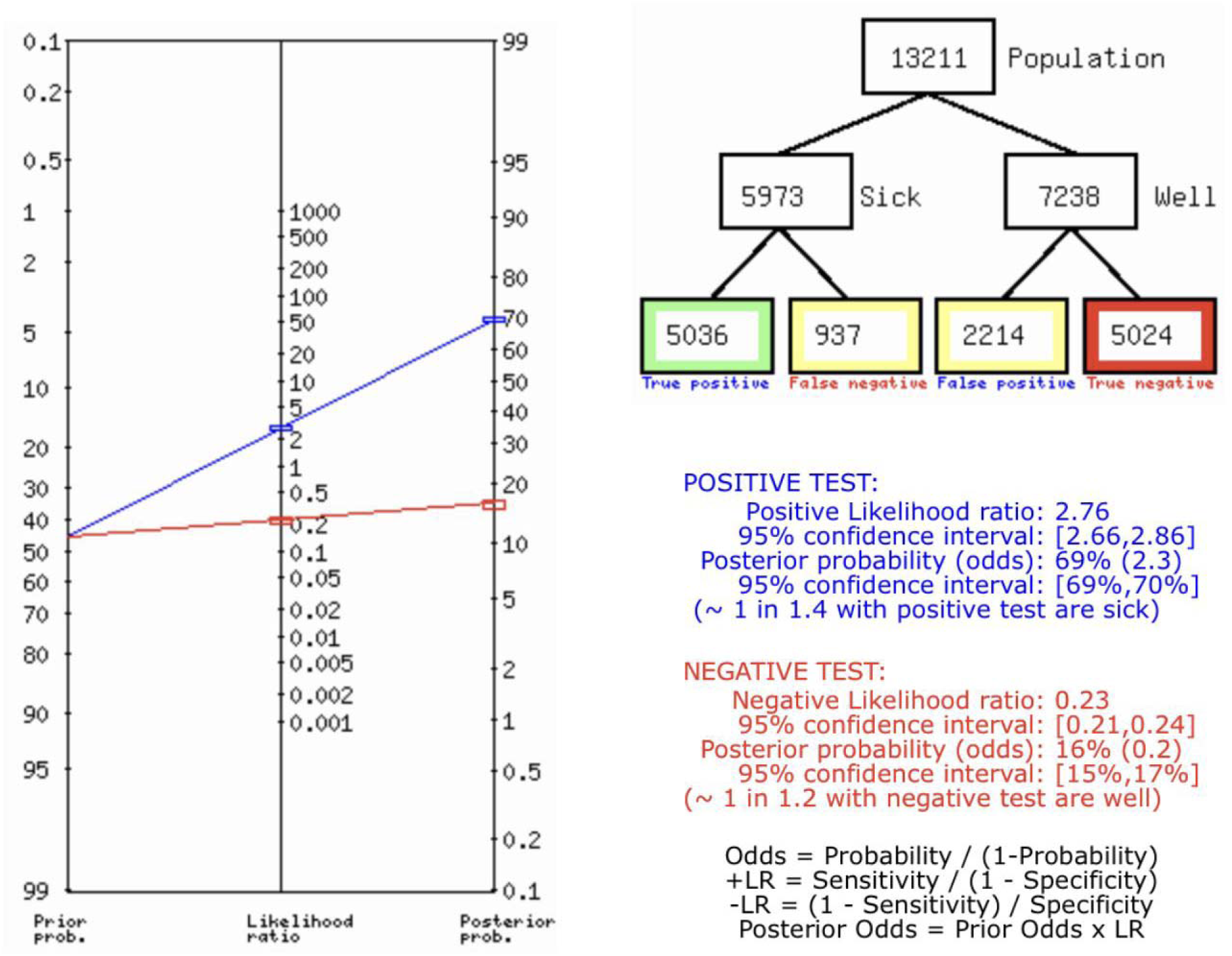
Fagan’s Analysis for Multiparametric MRI for Prostate Cancer Diagnosis.

<u>**Figure 7A describes the summary of Fagan plot analysis for all the studies considered for Multiparametric MRI, showing a prior probability of 45% (0.8); a Positive Likelihood Ratio of 2.76; a probability of post-test 69% (2.3); a Negative likelihood ratio of 0.23, and a probability of post-test 16% (0.2).**</u>

**Figure 7B :**
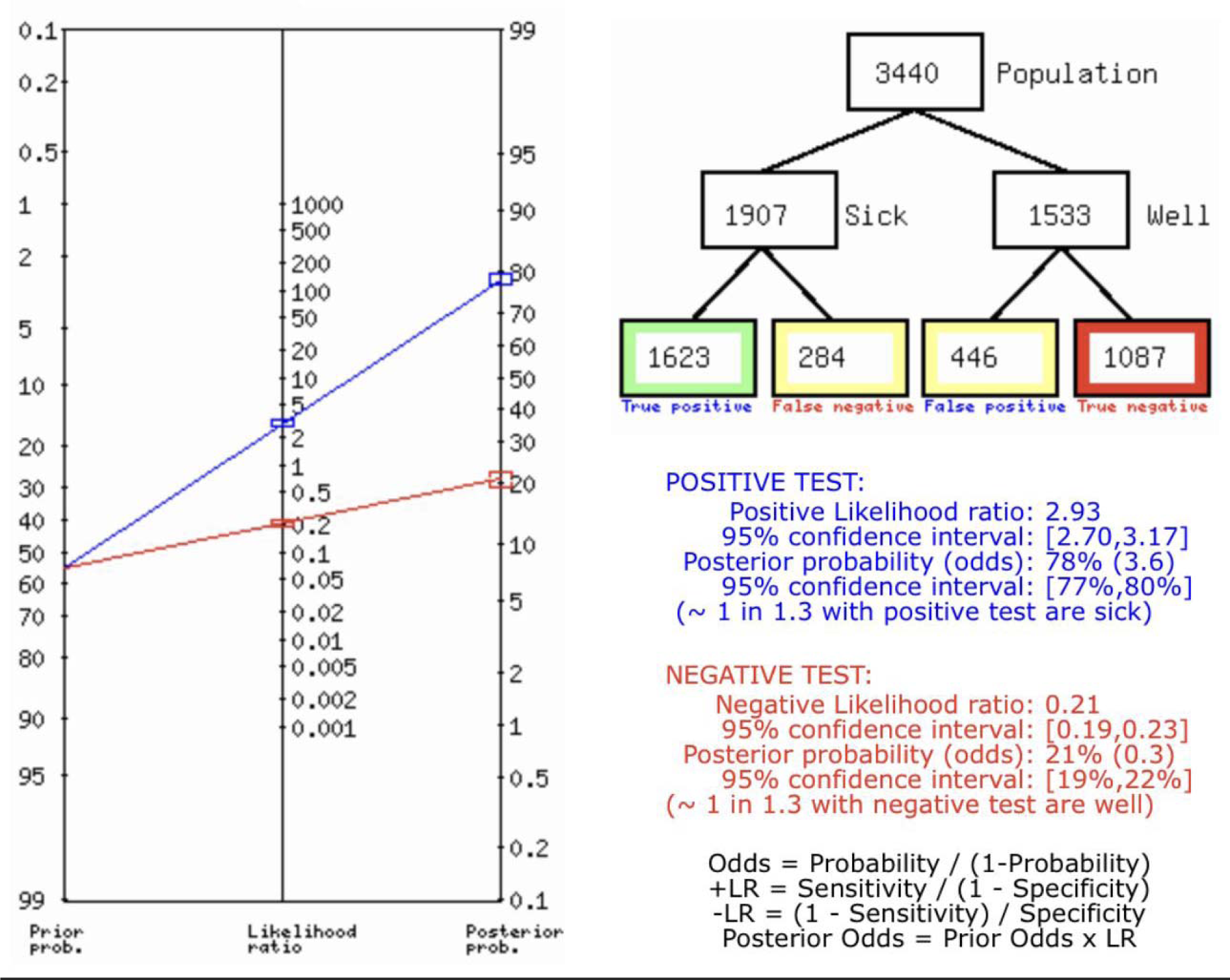
Fagan’s Analysis for Biparametric RI for Prostate Cancer Diagnosis.

<u>**Figure 7B describes the summary of Fagan plot analysis for all the studies considered for Biparametric MRI, showing a prior probability of 55% (1.2); a Positive Likelihood Ratio of 2.93; a probability of post-test 78% (3.6); a Negative likelihood ratio of 0.21, and a probability of post-test 21% (0.3).**</u>

## Bias study

### Bias study of multiparametric MRI

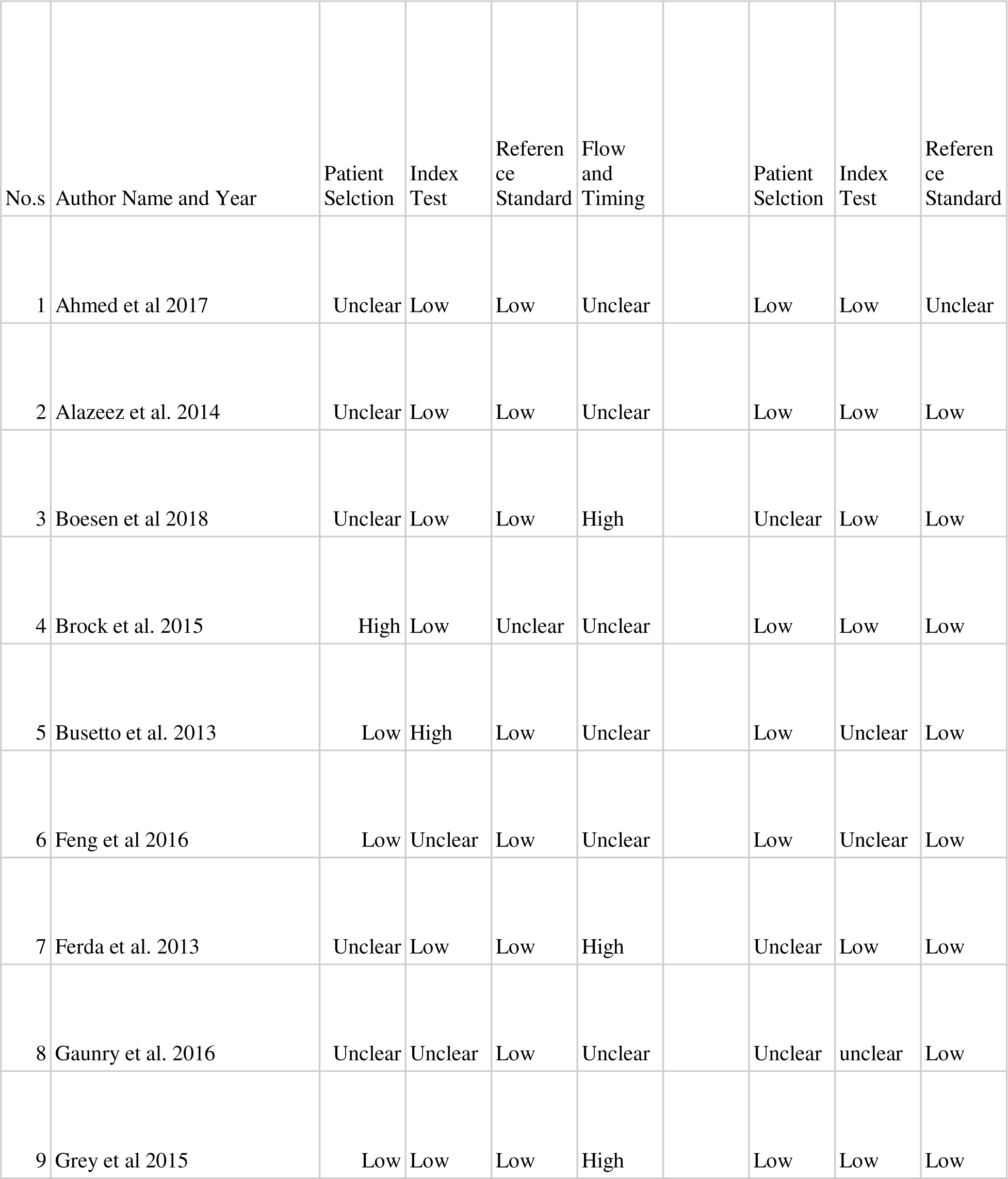
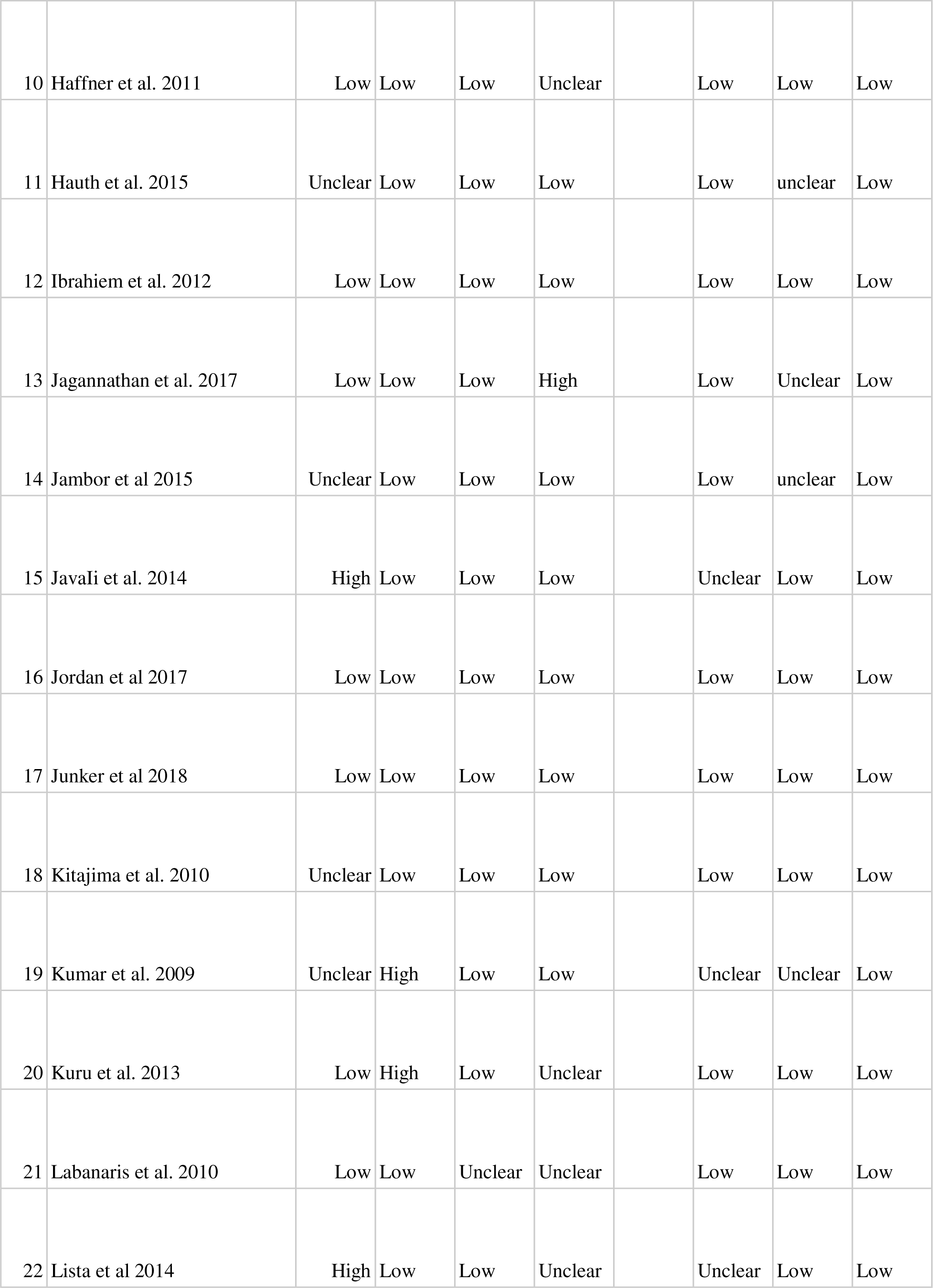
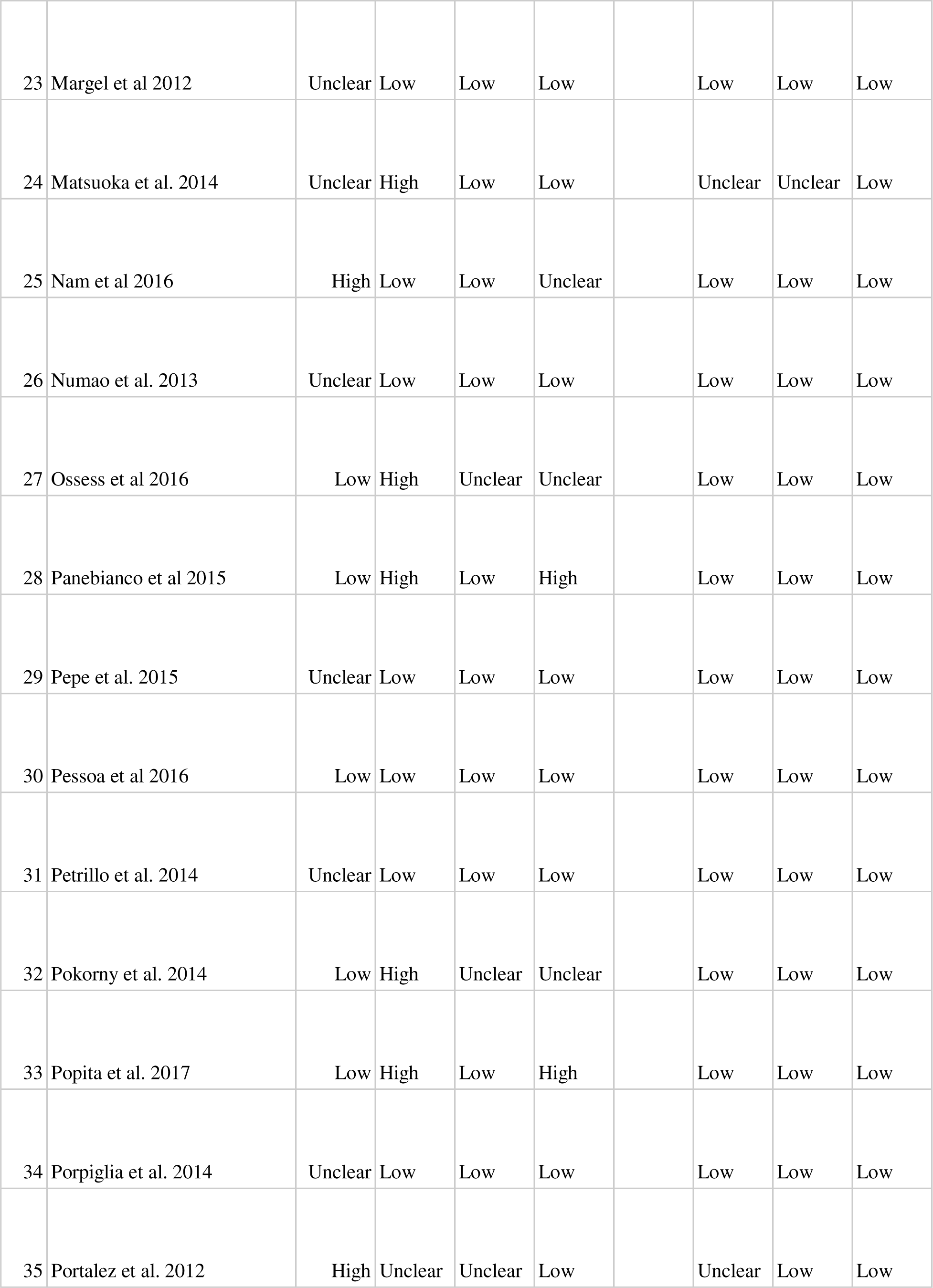
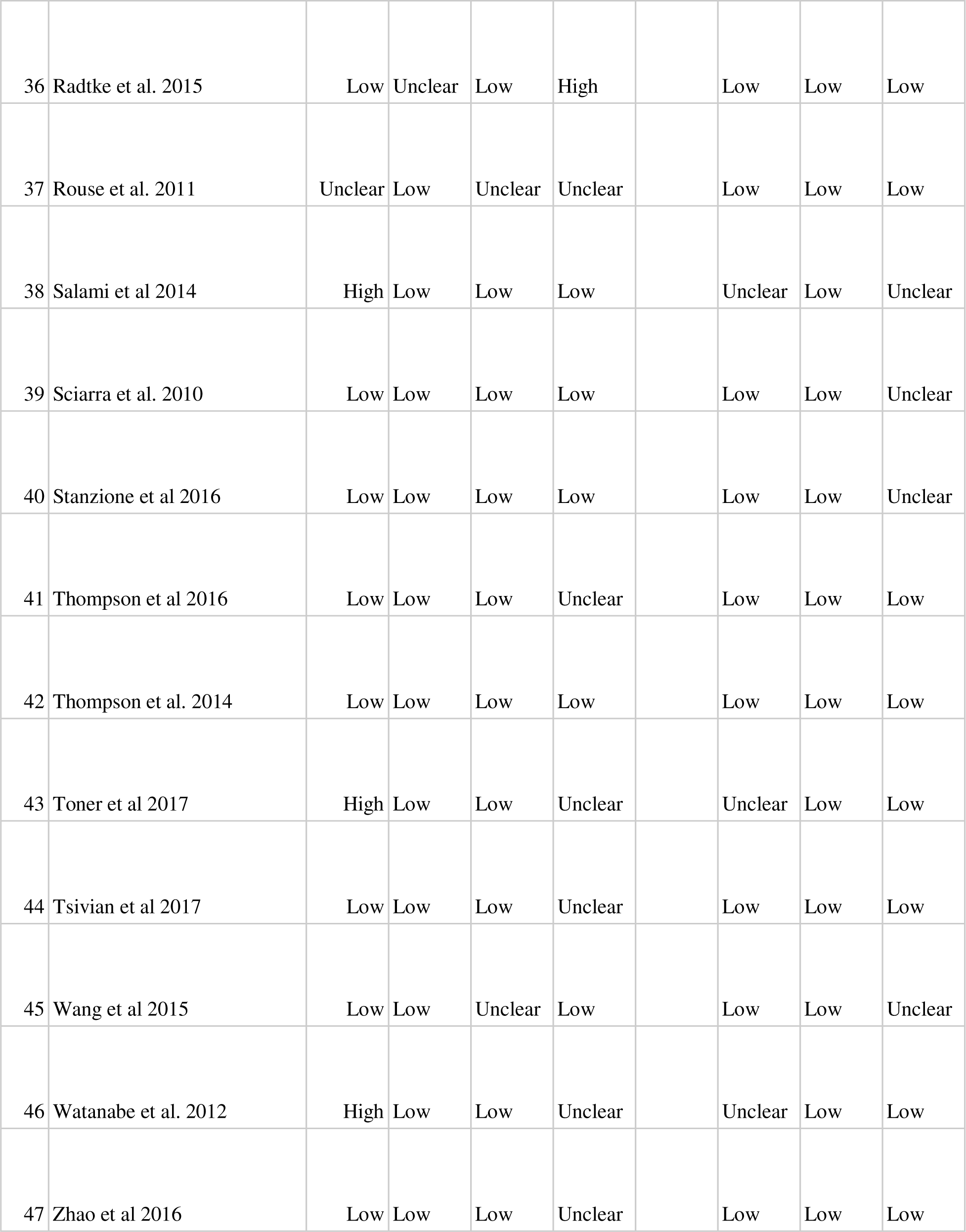
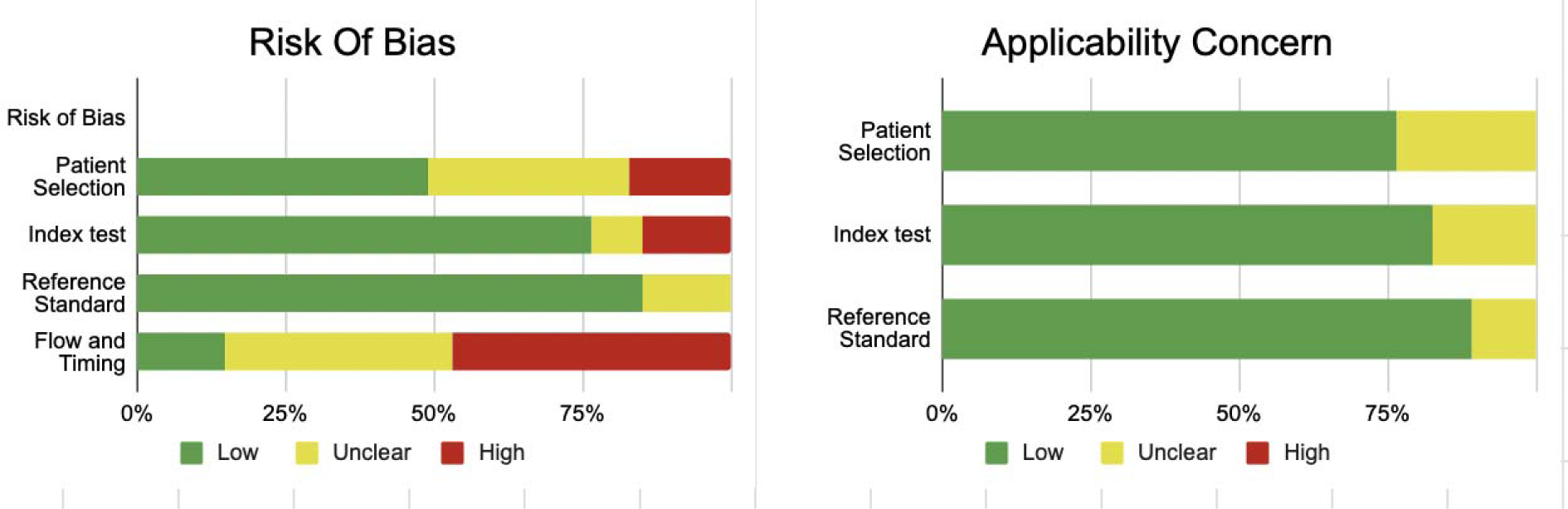

### Bias study of biparametric MRI

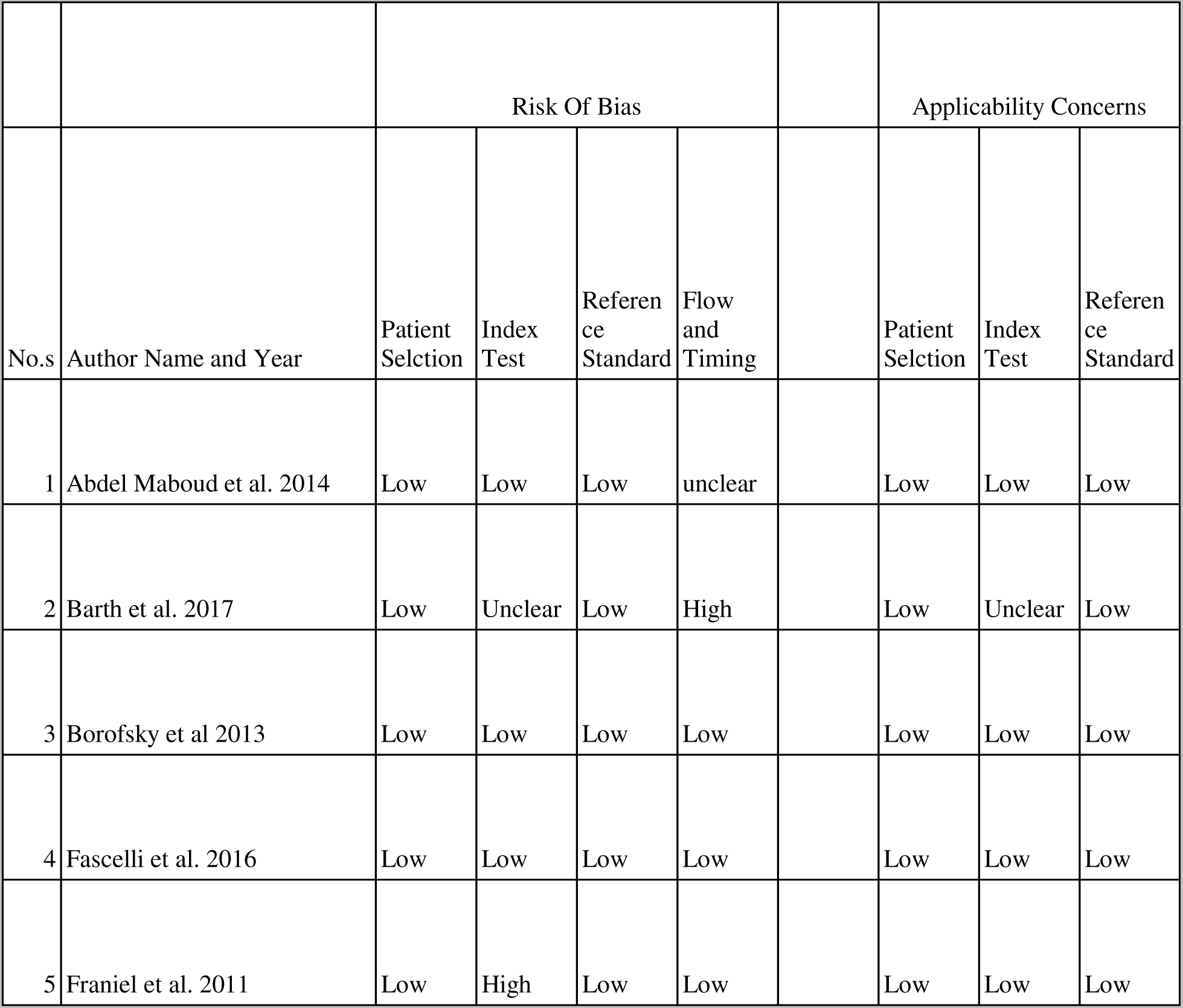
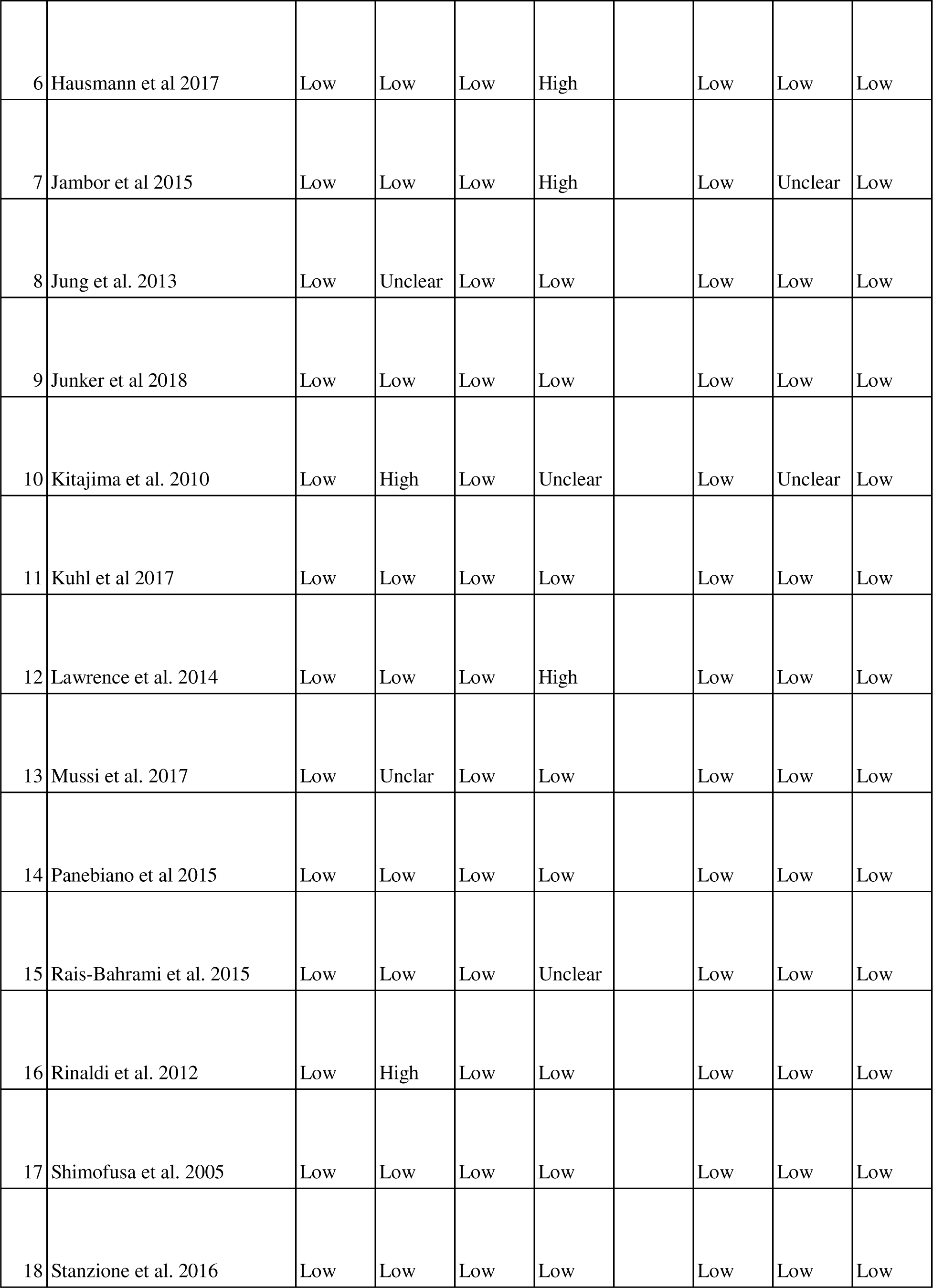
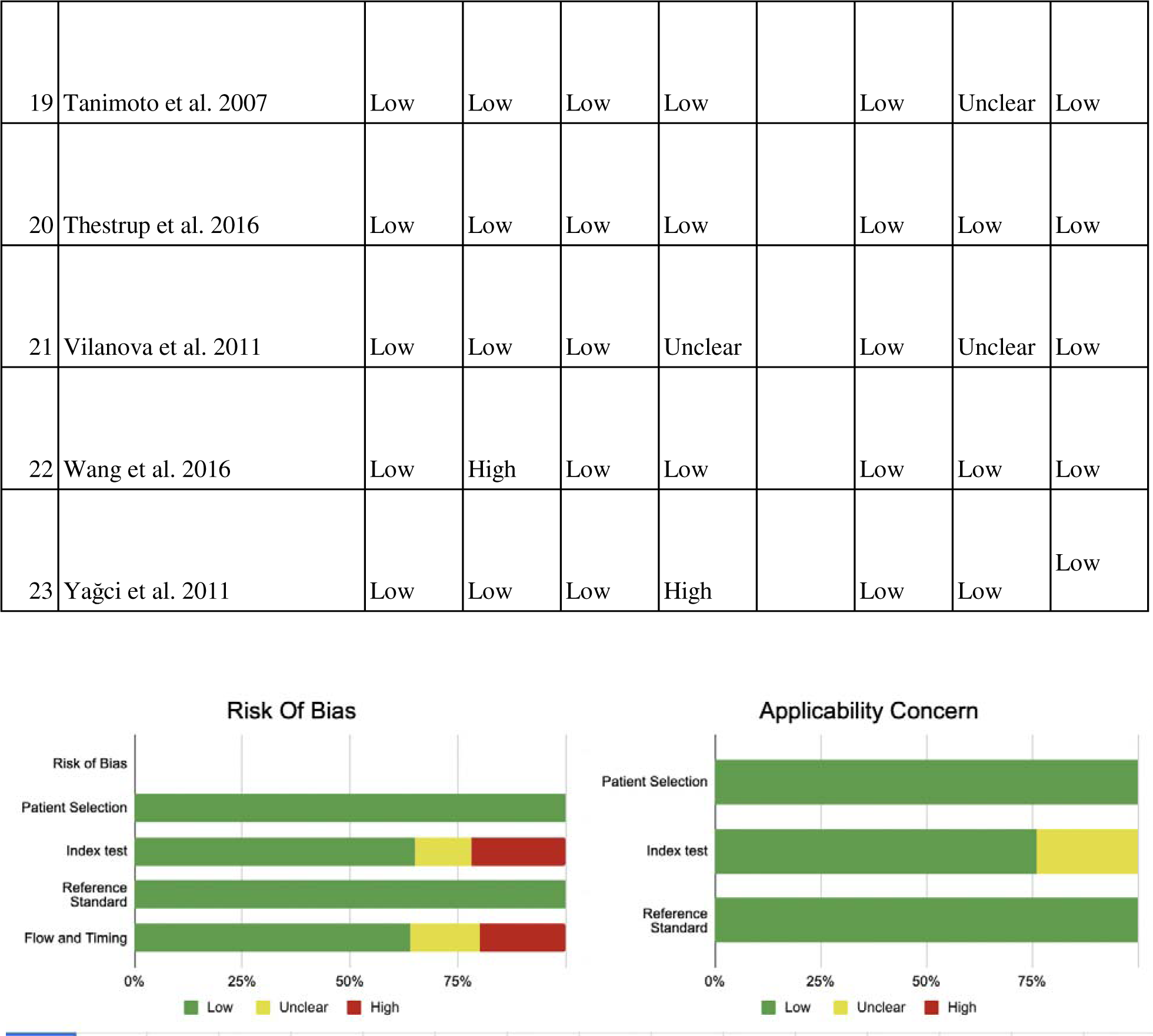

Bias study of biparametric MRI-
| No.s | Author Name and Year | Risk Of Bias |  |  |  | Applicability Concerns |  |  |
| --- | --- | --- | --- | --- | --- | --- | --- | --- |
|  |  | Patient Selection | Index Test | Reference Standard | Flow and Timing | Patient Selection | Index Test | Reference Standard |
| 1 | Abdel Maboud et al. 2014 | Low | Low | Low | unclear | Low | Low | Low |
| 2 | Barth et al. 2017 | Low | Unclear | Low | High | Low | Unclear | Low |
| 3 | Borofsky et al 2013 | Low | Low | Low | Low | Low | Low | Low |
| 4 | Fascelli et al. 2016 | Low | Low | Low | Low | Low | Low | Low |
| 5 | Franiel et al. 2011 | Low | High | Low | Low | Low | Low | Low |

|  |  |  |  |  |  |  |  |  |  |
| --- | --- | --- | --- | --- | --- | --- | --- | --- | --- |
| 6 | Hausmann et al 2017 | Low | Low | Low | High |  | Low | Low | Low |
| 7 | Jambor et al 2015 | Low | Low | Low | High |  | Low | Unclear | Low |
| 8 | Jung et al. 2013 | Low | Unclear | Low | Low |  | Low | Low | Low |
| 9 | Junker et al 2018 | Low | Low | Low | Low |  | Low | Low | Low |
| 10 | Kitajima et al. 2010 | Low | High | Low | Unclear |  | Low | Unclear | Low |
| 11 | Kuhl et al 2017 | Low | Low | Low | Low |  | Low | Low | Low |
| 12 | Lawrence et al. 2014 | Low | Low | Low | High |  | Low | Low | Low |
| 13 | Mussi et al. 2017 | Low | Unclear | Low | Low |  | Low | Low | Low |
| 14 | Panebianco et al 2015 | Low | Low | Low | Low |  | Low | Low | Low |
| 15 | Rais-Bahrami et al. 2015 | Low | Low | Low | Unclear |  | Low | Low | Low |
| 16 | Rinaldi et al. 2012 | Low | High | Low | Low |  | Low | Low | Low |
| 17 | Shimofusa et al. 2005 | Low | Low | Low | Low |  | Low | Low | Low |
| 18 | Stanzione et al. 2016 | Low | Low | Low | Low |  | Low | Low | Low |

|  |  |  |  |  |  |  |  |  |  |
| --- | --- | --- | --- | --- | --- | --- | --- | --- | --- |
| 19 | Tanimoto et al. 2007 | Low | Low | Low | Low |  | Low | Unclear | Low |
| 20 | Thestrup et al. 2016 | Low | Low | Low | Low |  | Low | Low | Low |
| 21 | Vilanova et al. 2011 | Low | Low | Low | Unclear |  | Low | Unclear | Low |
| 22 | Wang et al. 2016 | Low | High | Low | Low |  | Low | Low | Low |
| 23 | Yağci et al. 2011 | Low | Low | Low | High |  | Low | Low | Low |

### Publication bias

## Table of description

### Multiparametric MRI

Multiparametric MRI-
|  |  |  |  |  |  |  |  |
| --- | --- | --- | --- | --- | --- | --- | --- |
| Gaunry[7] | 2016 | 0.94 | 0.36 | 376 | 72 | 24 | 41 |
| Ahmed [8] | 2017 | 0.78 | 0.78 | 180 | 76 | 50 | 270 |
| Boesen[9] | 2018 | 0.92 | 0.75 | 81 | 50 | 7 | 151 |
| Brock[10] | 2015 | 0.89 | 0.16 | 56 | 88 | 7 | 17 |
| Busetto[11<br>] | 2013 | 0.9 | 0.62 | 61 | 36 | 7 | 59 |
| Feng [12] | 2016 | 0.96 | 0.84 | 144 | 40 | 6 | 211 |
| Ferda[13] | 2013 | 0.98 | 0.65 | 82 | 28 | 2 | 52 |
| Gaunry[7] | 2016 | 0.94 | 0.36 | 376 | 72 | 24 | 41 |
| Gaunay[7] | 2011 | 0.83 | 0.58 | 240 | 111 | 50 | 154 |
| Hauth[14] | 2015 | 0.98 | 0.12 | 42 | 45 | 1 | 6 |
| Ibrahiem[1<br>5] | 2012 | 0.84 | 0.58 | 57 | 10 | 11 | 14 |
| Jagannatha<br>n [15] | 2017 | 0.89 | 0.86 | 17 | 1 | 6 | 6 |
| Jambor<br>[16] | 2015 | 0.89 | 0.5 | 28 | 11 | 3 | 11 |
| JavaIi[17] | 2014 | 0.96 | 0.42 | 22 | 68 | 1 | 49 |
| Jordan[18]<br>, [19] | 2017 | 0.77 | 0.59 | 63 | 82 | 18 | 119 |
| Junker[20] | 2018 | 0.98 | 0.45 | 133 | 55 | 2 | 46 |
| Kitajima[2<br>1] | 2010 | 0.81 | 0.96 | 80 | 14 | 19 | 311 |
| Kumar<br>[22] | 2009 | 0.77 | 0.83 | 10 | 8 | 3 | 39 |
| Kuru [23] | 2013 | 0.83 | 0.3 | 67 | 186 | 14 | 80 |
| Labanaris[<br>24] | 2010 | 0.57 | 0.19 | 96 | 74 | 73 | 17 |
| Lista [25],<br>[26] | 2014 | 0.81 | 0.65 | 23 | 42 | 5 | 80 |
| Margel<br>[26] | 2012 | 0.55 | 0.94 | 10 | 2 | 8 | 36 |
| Matsuoka<br>[27] | 2014 | 0.75 | 0.64 | 149 | 26 | 49 | 46 |
| Nam [28] | 2016 | 0.65 | 0.84 | 6 | 3 | 3 | 18 |
| Numao[27<br>] | 2013 | 0.64 | 0.71 | 101 | 56 | 57 | 136 |
| Ossess[29] | 2015 | 0.98 | 0.38 | 62 | 57 | 1 | 36 |
| Panebianco [30] | 2015 | 0.91 | 0.71 | 642 | 63 | 63 | 157 |
| Pepe[31] | 2015 | 0.89 | 0.86 | 49 | 4 | 6 | 24 |
| Pessoa[32] | 2016 | 0.92 | 0.8 | 54 | 9 | 4 | 38 |
| Petrillo [33] | 2014 | 0.84 | 0.5 | 21 | 55 | 4 | 56 |
| Pokorny[34] | 2014 | 0.8 | 1 | 101 | 0 | 25 | 56 |
| Popita [35] | 2017 | 1 | 0.83 | 16 | 4 | 0 | 19 |
| Porpiglia[36] | 2014 | 0.9 | 0.91 | 47 | 11 | 5 | 107 |
| Portalez [37] | 2012 | 0.42 | 0.9 | 34 | 47 | 47 | 404 |
| Radtke[23] | 2015 | 0.48 | 0.96 | 72 | 6 | 78 | 138 |
| Rouse [38] | 2011 | 0.87 | 0.67 | 74 | 72 | 11 | 145 |
| Salami [39] | 2014 | 0.78 | 0.69 | 65 | 28 | 18 | 64 |
| Sciarra [40] | 2010 | 0.94 | 0.87 | 66 | 9 | 4 | 61 |
| Stanzione [41] | 2016 |  |  | 31 | 2 | 3 | 51 |
| Thompson [42] | 2016 | 0.90 | 0.95 | 96 | 28 | 45 | 175 |
| Thompson [43] | 2014 | 0.96 | 0.37 | 137 | 127 | 6 | 74 |
| Toner[44] | 2017 | 0.63 | 0.5 | 58 | 30 | 34 | 30 |
| Tsivian[45] | 2017 | 0.93 | 0.44 | 32 | 8 | 2 | 8 |
| Wang[46] | 2015 | 0.9 | 0.91 | 306 | 60 | 34 | 613 |
| Watanabe[47] | 2012 | 0.9 | 0.65 | 624 | 266 | 73 | 485 |
| Zhao [48] | 2016 | 0.88 | 0.84 | 137 | 34 | 18 | 183 |

### Biparametric MRI

**Biparametric MRI-**
| **Author** | **year** | **sensitivity** | **specificity** | **True Positive (Tp)** | **False Positive (FP)** | **False Negative (FN)** | **True Negative (TN)** |
| --- | --- | --- | --- | --- | --- | --- | --- |
| [49] | 2014 | 0.87 | 0.8 | 27 | 1 | 4 | 4 |
| Barth[50] | 2017 | 0.95 | 0.9 | 37 | 2 | 1 | 23 |
| Borofsky [51] | 2013 | 0.65 | 0.65 | 56 | 24 | 29 | 45 |
| Fascelli[52] | 2016 | 0.91 | 0.53 | 41 | 7 | 3 | 8 |
| Franiel [53] | 2011 | 0.85 | 0.37 | 18 | 20 | 3 | 13 |
| Hausmann[54] | 2017 | 0.87 | 0.84 | 54 | 4 | 8 | 27 |
| Jambor [55] | 2015 | 0.71 | 0.73 | 23 | 5 | 8 | 17 |
| Jung [56] | 2013 | 0.68 | 0.7 | 49 | 25 | 23 | 59 |
| Junker[57] | 2018 | 0.99 | 0.39 | 134 | 61 | 1 | 40 |
| Kitajima [58] | 2010 | 0.76 | 0.94 | 23 | 1 | 7 | 22 |
| Kuhl[59] | 2017 | 0.94 | 0.88 | 139 | 47 | 8 | 348 |
| Lawrence [60] | 2014 | 0.84 | 0.66 | 14 | 7 | 2 | 16 |
| Mussi [61] | 2017 | 0.52 | 0.79 | 36 | 10 | 32 | 40 |
| Panebianco [62] | 2015 | 0.87 | 0.85 | 614 | 33 | 91 | 187 |
| Rais-Bahrami [63] | 2015 | 1 | 0.12 | 84 | 51 | 0 | 8 |
| Rinaldi [64] | 2012 | 0.87 | 0.67 | 32 | 1 | 4 | 4 |
| Shimofusa [65] | 2002 | 0.86 | 0.83 | 32 | 3 | 5 | 20 |
| Stanzione[66] | 2016 | 0.85 | 0.98 | 29 | 0 | 5 | 48 |
| Tanimoto [67] | 2007 | 0.84 | 0.85 | 37 | 5 | 7 | 34 |
| Thestrup [68] | 2016 | 0.94 | 0.15 | 64 | 115 | 4 | 21 |
| Vilanova [69] | 2011 | 0.74 | 0.91 | 29 | 2 | 9 | 30 |
| Wang [46] | 2016 | 0.54 | 0.89 | 33 | 8 | 27 | 65 |
| Yağci [70] | 2011 | 0.81 | 0.32 | 18 | 14 | 3 | 8 |

## Discussion

Prostate cancer is a prevalent malignancy that primarily affects the prostate gland in men. It typically develops slowly and may not cause noticeable symptoms in its early stages. Early detection through screening and timely intervention can significantly improve outcomes for individuals with prostate cancer.

Prostate cancer can be diagnosed through the PSA blood test, which measures prostate-specific antigen levels. Digital rectal examinations and prostate biopsies are also used to assess and confirm the presence of cancer, with biopsies offering a definitive diagnosis. However, these methods can be uncomfortable, carry a risk of complications and may cause physical and psychological distress to patients.

Among the non invasive methods, MRI has gained popularity is diagnosis of prostate cancer. Early and prompt diagnosis plays a vital role in ensuring a proper course of action to deal with the disease process.

This meta-analyis aims to compare the diagnostic accuracy between multiparametric and biparametric MRI for diagnosis of prostate cancer.

Multiparametric-MRI combines one or more functional MRI techniques, such as diffusion-weighted imaging (DWI) and dynamic contrast enhanced (DCE) imaging, with high-resolution anatomical T2-weighted (T2W) and T1-weighted (T1W) images.[9]

It offers comprehensive insights into prostate anatomy, diffusion patterns, vascularity, and function, making it the preferred gold standard for prostate imaging. MpMRI finds extensive clinical use in detecting prostate cancer, pinpointing its location, assessing risk levels, guiding treatment decisions, and monitoring disease progression.

For the analysis, a total of 47 RCTs with 13,211 participants were chosen. Of these, 8 studies demonstrated sensitivity above or equal to 95% and 5 studies demonstrated specificity over 95%.

Multiparametric MRI has a sensitivity of 0.84 and specificity of 0.69, with a 95% confidence interval falling between 0.68 and 0.70.

Biparametric MRI is a simplified version of multiparametric MRI and relies on T2-weighted and DWI sequences. The Brownian motion of free water in tissue is evaluated by DWI. Water motion is constrained in tumours, possibly as a result of their higher cellular density and increased nucleocytoplasmic ratio, and it can be shown on ADC maps, allowing for a quantitative assessment.[70]

The study analyzed 23 randomized controlled trials with 3,440 participants. Four trials had sensitivity rates of 95% or higher, while two had specificity rates above 95%. Biparametric MRI had a sensitivity of 0.85 and specificity of 0.71 (CI: 0.69-0.73).

However, in comparison to mpMRI, the time needed for an MRI with bpMRI without DCE-MRI is shorter, the cost-effectiveness for patients is improved, and there is less fear about side effects.

PI-RADS (Prostate Imaging Reporting and Data System) aims to standardize prostate mpMRI procedures, interpretation, and reporting for better prostate cancer patient management. PI-RADS V2 introduced a key change by emphasizing a dominant sequence approach: T2-weighted imaging (T2WI) for transition zone lesions and diffusion-weighted imaging (DWI) for peripheral zone lesions, distinguishing it from the 2012 version[71].

Keeping into consideration the pros and cons of both forms of MRI, choice is made depending on resource availability, clinical goals, and patient-specific needs. MpMRI is the preferred option because of its improved accuracy and thorough evaluation, especially in high-tech healthcare settings. Nevertheless, bpMRI can be a workable substitute in circumstances where a more simplified technique is required.

## Data Availability

All data produced in the present work are contained in the manuscript

